# Predicting cohort-specific cervical cancer incidence from population-based HPV prevalence surveys: a worldwide study

**DOI:** 10.1101/2020.03.10.20031013

**Authors:** Rosa Schulte-Frohlinde, Damien Georges, Gary M. Clifford, Iacopo Baussano

**Affiliations:** International Agency for Research on Cancer, Lyon, France; Department of Virology, University of Leipzig, Leipzig, Germany

**Keywords:** Cervical cancer, HPV infection, risk prediction

## Abstract

**Background:** Predictions of cervical cancer burden and impact of control measures are often modelled from HPV prevalence. However, predictions could be improved by data on time between prevalent HPV detection and cervical cancer occurrence.

**Methods:** Based upon high-risk (HR) HPV prevalence and cervical cancer incidence in the same birth cohorts from 17 worldwide locations, and informed by individual-level data on age at HR HPV detection and on sexual debut, we built a mixed model to predict cervical cancer incidence up to 14 years following prevalent HR HPV detection.

**Findings:** Cervical cancer incidence increased significantly during the 14 years following HR HPV detection in women <35 years, e.g. from 0·02 (95% CI 0·003–0·06) per 1000 within 1 year to 2·8 (1·2–6·5) at 14 years for unscreened women, but remained relatively constant following prevalent HR HPV detection above 35 years, e.g. from 5·4 (2·5–11) per 1000 within 1 year to 6·4 (2·4–17·1) at 14 years for unscreened HR HPV positive women aged 45–54 years. Age at sexual debut was a significant modifier of cervical cancer incidence in HR HPV positive women aged <25, but less so at older ages, whereas screening was a modifier in women ≥35 years. Lastly, we predicted annual number and incidence of cervical cancer in ten additional IARC HPV prevalence survey locations without representative cancer incidence data.

**Interpretation:** These findings can inform cervical cancer control programmes, particularly in settings without cancer registries, as they allow prediction of future cervical cancer burden from population-based surveys of HPV prevalence.

**Funding:** Bill & Melinda Gates Foundation; Canadian Institutes of Health Research.

## Introduction

Despite being largely preventable, cervical cancer is a leading cause of morbidity and mortality among women worldwide with approximately 570 000 new cases and 310 000 deaths in 2018.^1^ Nearly half of the cases are diagnosed in women younger than 50 years of age and more than two-thirds occur in low and middle-income countries (LMICs).^2^

Persistent infection with one of 13 high-risk (HR) human papillomavirus (HPV) types is the necessary cause of cervical cancer.^3^ HPV16 and 18, targeted by all licensed vaccines, are responsible for 70% of cervical cancer; whereas HPV types 31/33/45/52/58 targeted by the nonavalent vaccine are responsible for another 18%.^4^ HPV vaccines have demonstrated high safety^5,6^ and efficacy against persistent HPV infections, pre-cancerous lesions.^7^ Recent data also suggest high efficacy against invasive cervical cancers.^8^ HPV vaccination programs have been shown to be cost-effective in a wide range of settings and conditions.^9^ Nevertheless, HPV vaccine has been introduced mostly in high-resource settings, whereas access to HPV vaccination remains limited especially in LMICs, in particular Africa and Asia.^10^ Similarly, the coverage of cervical cancer screening, which has been shown to be effective in controlling the burden of cervical cancer in high-income countries (HICs), is highly heterogeneous worldwide and remains very low in LMICs.^11^ HPV vaccination, cervical cancer screening, and the management of detected disease are the main components of the recent strategy launched by WHO to eliminate cervical cancer as a public health problem.^12^

An accurate quantification of cervical cancer burden is essential to inform the planning of cancer control programmes and to monitor the impact of control measures. The most valid estimates of cancer burden are obtained from population-based cancer registries, which are, however, resource demanding. On a global scale, about a third of countries (65 countries) have high quality national (or subnational) data on cancer incidence,^13^ but this coverage, although improving in LMICs, remains mostly confined to HICs. Hence, present and future trends of cervical cancer incidence in most LMIC remain, and will remain, unknown.

Both mathematical^14^ and statistical^15^ models are extensively used to predict the burden of cervical cancer and to project the impact of control measures in absence of local high quality data. Nevertheless, the average time between HPV infection acquisition and cervical cancer occurrence remains difficult to assess, firstly because it is far more feasible to have data on prevalence rather than on incidence of HPV, and also because follow-up of pre-cancerous lesions without treatment to understand their cervical cancer incidence is unethical.^16^ This uncertainty may affect the validity of the cervical cancer risk estimates and of the expected impact of preventive measures in specific populations.

To partially overcome this limitation, we have conducted a birth cohort-specific ecological study accounting for time-lag between HR HPV prevalence measurement and cervical cancer detection, hereafter labelled as “time-lag” for short, to describe the age-specific association between HR HPV prevalence and cervical cancer incidence in the same birth cohorts and to estimate cervical cancer incidence rates in HR HPV positive women.

## Methods

### Data sources

Age-stratified and population-based HR HPV prevalence data were obtained from the International Agency for Research on Cancer (IARC) HPV Prevalence Surveys database (appendix p 2, Fig S1). All cross-sectional surveys (n=28) used a standardized protocol for population-based recruitment and detection of HPV in cervical cell samples using a GP5+/6+-based PCR assay detecting at least 36 types.^17^ The following HPV types were defined as HR types: 16, 18, 31, 33, 35, 39, 45, 51, 52, 56, 58, 59, and 68. A woman was excluded from the prevalence estimate if known to be currently pregnant or hysterectomised, if data on age or HPV status was missing or if her cervical specimen was beta-globulin negative.

Cervical cancer incidence data were extracted from cancer registries included in Cancer Incidence in Five Continents (CI5), Volume VIII-XI, a series of monographs published every five years by IARC and the International Association of Cancer Registries (IACR) including age-stratified data from high quality cancer registries worldwide.^13,18-20^

### Association between HR HPV prevalence and cervical cancer incidence

HR HPV prevalence and cervical cancer incidence data in the same birth cohorts were available for 17 locations (appendix p 2, Fig S1 and appendix p 7, Table S1) and were used to describe the age-specific effect of HR HPV prevalence on cervical cancer incidence. For 12 locations, HR HPV prevalence survey data and incidence data from cancer registry were available from the same geographic area. For the prevalence studies in Warsaw, Poland; Barcelona, Spain; Bogota, Colombia; Shanxi, China; and Tehran, Iran, we selected cancer registries, with comparable characteristics whenever possible, in the same region or country, namely: Kielce, showing similarly low rates as Warsaw in previous years; Tarragona, the next largest city after Barcelona in the region of Catalonia; Cali, the largest Colombian cancer registry; Cixian, as the study in Shanxi also included women from this region; and Gholestan province in Iran.

We stratified our populations in 10-year age groups (25-34, 35-44, 45-54, and 55-64-year-old at HPV detection) and assessed the association between age-specific HR HPV prevalence and cervical cancer incidence for the same 10-year birth cohort fitting an additive Poisson regression model (without an intercept term to comply with the criteria that cervical cancer rates should be zero if the HPV prevalence is zero). In the regression model, we also included age at HPV detection and location-specific time-lag (time between assessment of HR HPV prevalence and cancer incidence in the same 10-year birth cohort; <10 or ≥10 years) as modifiers of the effect of HPV prevalence on cervical cancer incidence rates. Since cervical cancer screening is an important confounding factor at a population level, we also conducted a sensitivity analysis excluding data from locations with organized screening programmes with reported coverage above 50%, i.e. Amsterdam, the Netherlands and Turin, Italy, hereafter labelled as “locations with screening”.^21^ The other locations are considered as “locations without screening” for the purpose of the present analysis.^11,22^

### Cervical cancer incidence among HR HPV positive women

In this analysis, we included a subset of 16 locations (appendix p 7, Table S1) for which cervical cancer incidence data from CI5 Volumes VIII-XI covered relevant time intervals, to calculate country-standardized and cohort-specific cervical cancer incidence rates in HR HPV positive women (only Ho Chi Minh City, Vietnam, was excluded). We stratified our populations in the age groups 20-24, 25-34, 35-44, and 45-54 year-old at HPV detection. For each location and year since HPV measurement, we calculated the cohort-specific number of HR HPV positive women and cervical cancers, and computed the cohort-specific incidence of cervical cancer in HR HPV positive women (see appendix p 2, Figure S1 for details).

To estimate cohort-specific incidence of cervical cancer in women with prevalent HR HPV infection, we fitted, for each age-group at HPV detection, a mixed effect linear regression model with years since HPV detection, average age at sexual debut, their interaction, and screening implementation status as covariates, location as grouping variable, and the risk of cervical cancer (on a log scale) as an outcome. To be able to include at least three locations in each analysis, we limited the time interval for cancer incidence estimates to 14 years. For the same 14 locations without screening, average age at sexual debut (appendix p 8, Table S2) was estimated from the IARC HPV Prevalence Surveys database,^17^ whereas for Amsterdam and Turin it was obtained from national population-based surveys.^23,24^ To minimize the impact on our findings of HPV prevalence estimates based on few HPV infections, we also performed a sensitivity analysis restricted to 14 locations in which HPV prevalence was estimated on at least ten infections in each age group (i.e. we excluded Barcelona, Spain, and Songkla, Thailand). Based on mixed effect model-based estimates, we have drawn cervical cancer incidence predictions for specific ages at sexual debut and, based on the fixed effect component of the regression models, we predicted the incidence in each location. We used bootstrap for mixed models to calculate 95% prediction intervals (PIs) of cervical cancer incidence.

Finally, to exemplify how the proposed model can be used to predict cervical cancer incidence from estimates of age-specific HR HPV prevalence and average age at sexual debut, we used data from IARC population-based HPV surveys not included in the previous analysis (due to unavailability of cancer incidence data from CI5) to calculate the expected annual cervical cancer burden (numbers and incidence) in the population of the corresponding country ten years after the implementation of the surveys.

### Role of the funding source

The funder of the study had no role in study design, data collection, data analysis, data interpretation, or writing of the report. The corresponding author had full access to all the data in the study and had final responsibility for the decision to submit for publication.

## Results

Fig 1 shows, for 17 locations, cervical cancer incidence rates per 100 000 women plotted against HR HPV prevalence, by age group, as well as the estimated effect of HR HPV prevalence on cervical cancer incidence rates. Estimated cervical cancer incidence increased with HR HPV prevalence in all age groups. The size of the effect of HPV prevalence on cancer incidence rates increased with age and was also systematically higher in locations with a longer time-lag between HPV and cervical cancer assessment in the same birth cohorts (Fig 1). The effect ranged from 0·83 (0·75–0·9) to 2·3 (2·2–2·4) for women aged 25–34 years at HR HPV detection and from 4·6 (4·3–5·0) to 7·4 (7·0–7·8) for women aged 55–64 years, for a time-lag of <10 and ≥10 years, respectively (Table 1). Effect estimates were not materially different in a sensitivity analysis excluding locations with screening (appendix p 9, Table S3).

**Table 1.** Estimated effect of HR HPV prevalence on cervical cancer incidence rates by age at HPV detection and time-lag between HPV prevalence and cancer incidence measurement.

| Time-lag<br>(years)* | Age group at HPV detection (years) |  |  |  |
| --- | --- | --- | --- | --- |
|  | 25–34 | 35–44 | 45–54 | 55–64 |
| <10 | 0.83 (0.75–0.9) | 2.4 (2.3–2.6) | 3.5 (3.3–3.7) | 4.6 (4.3–5) |
| ≥10 | 2.3 (2.2–2.4) | 5.8 (5.6–6) | 6 (5.7–6.3) | 7.4 (7–7.8) |

\*Years elapsed between HR HPV detection and cervical cancer incidence assessment in the same birth cohorts. HR HPV = high-risk human papillomavirus.

**Fig 1.**
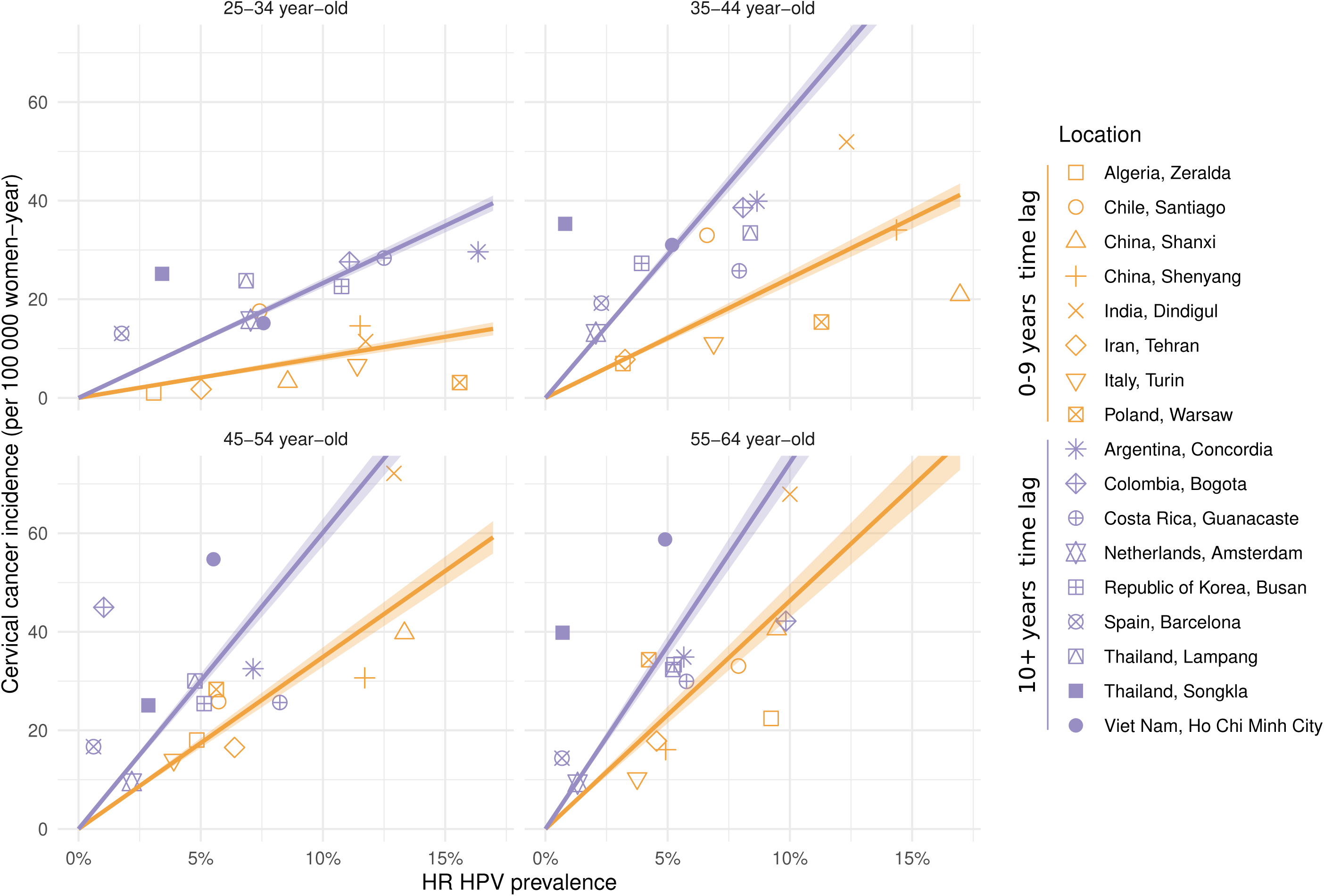
Birth cohort-specific HR HPV prevalence and cervical cancer incidence rates, by age-group at HPV detection, time-lag between HR HPV prevalence measurement and cervical cancer detection, and location. Colored lines represent effect estimates (with 95% confidence intervals) obtained fitting a Poisson regression model. HR HPV=high-risk human papillomavirus.

Results of the mixed effect regression model are shown in Table 2. Time-lag between HR HPV prevalence and cancer incidence assessment significantly affected cervical cancer risk in women below 35 years of age, and was also significantly modified by the average age at sexual debut. Above age 35, however, cervical cancer risk was not affected by either variable. The intraclass correlation coefficient, i.e. the proportion of the variance explained by setting location as grouping variable, increased with age from 0·6 in 20 to 24 year-olds to more than 0·9 in 45 to 54 year-olds. Similar results were obtained analysing the dataset restricted to locations with at least ten HPV infections in each age group (appendix pp 10-11, Table S4).

**Table 2.**
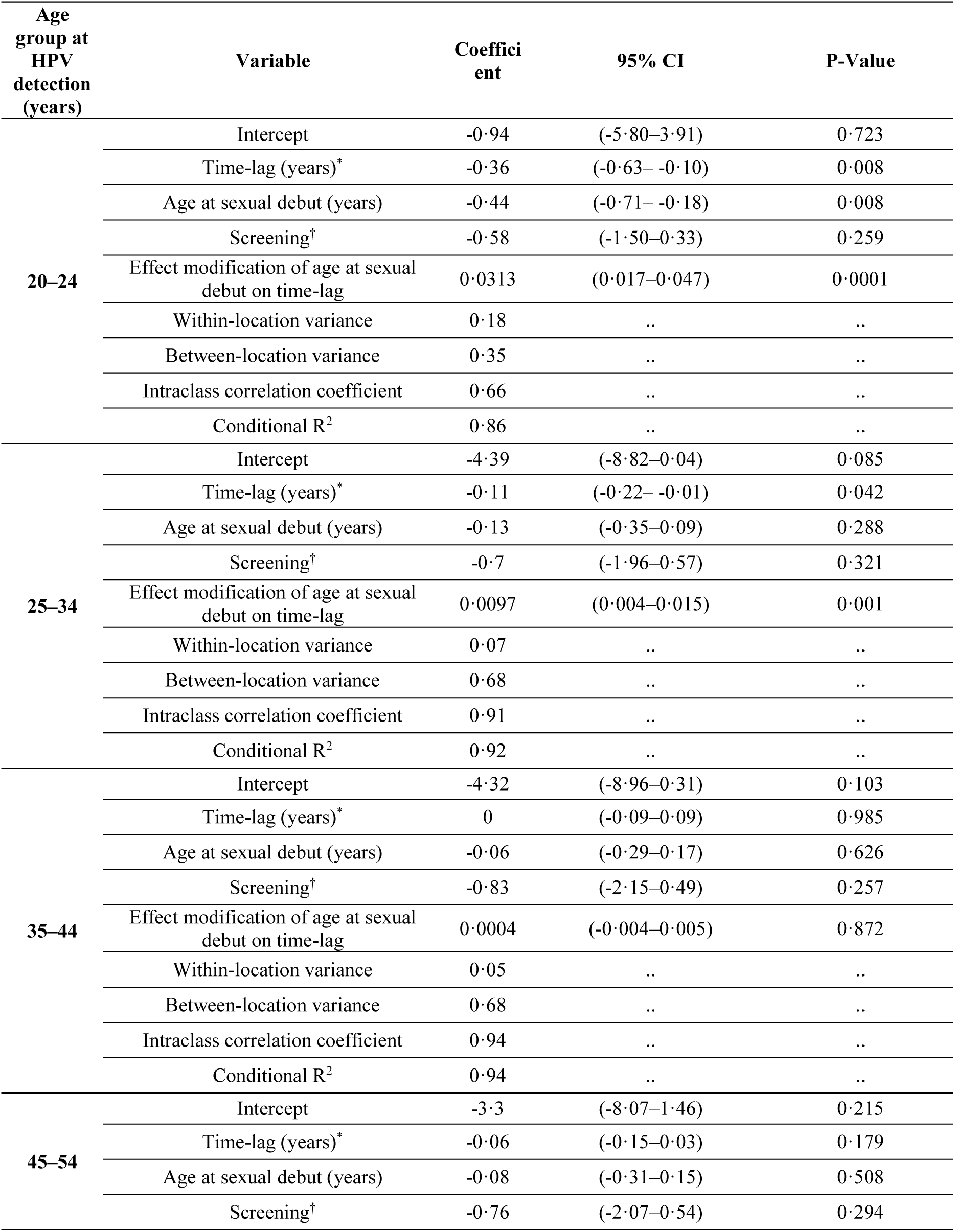
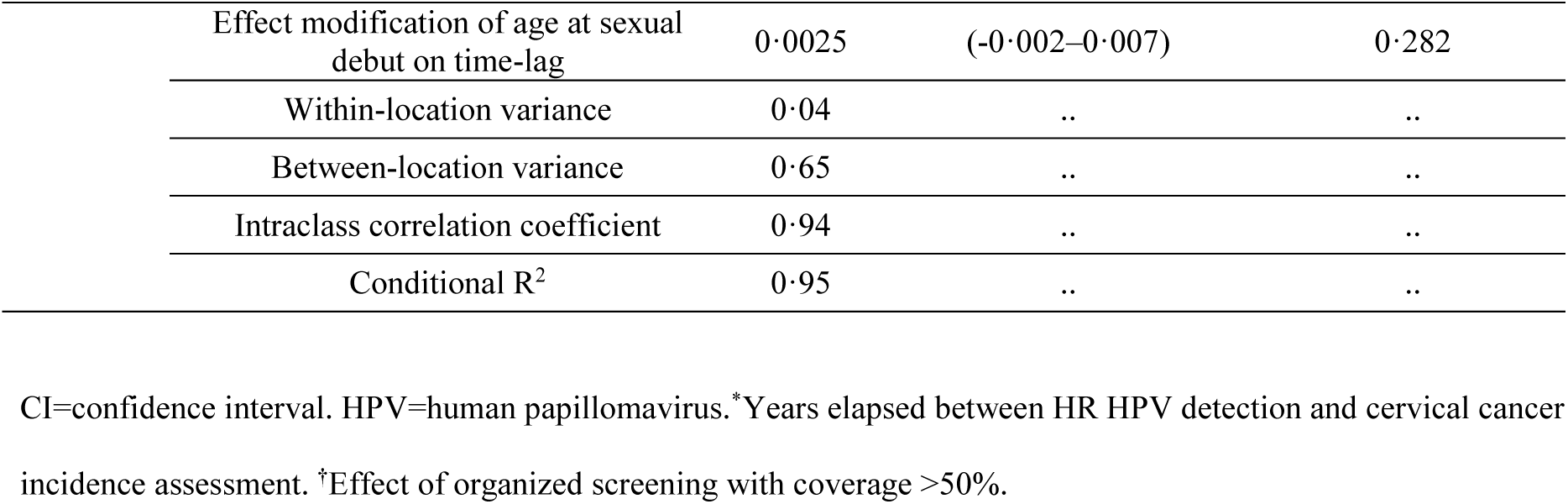
Results of the mixed effect linear regression models, by age group at HPV detection.

| Age group at HPV detection (years) | Variable | Coefficient | 95% CI | P-Value |
| --- | --- | --- | --- | --- |
| 20–24 | Intercept | -0.94 | (-5.80–3.91) | 0.723 |
|  | Time-lag (years)* | -0.36 | (-0.63– -0.10) | 0.008 |
|  | Age at sexual debut (years) | -0.44 | (-0.71– -0.18) | 0.008 |
|  | Screening<sup>†</sup> | -0.58 | (-1.50–0.33) | 0.259 |
|  | Effect modification of age at sexual debut on time-lag | 0.0313 | (0.017–0.047) | 0.0001 |
|  | Within-location variance | 0.18 | .. | .. |
|  | Between-location variance | 0.35 | .. | .. |
|  | Intraclass correlation coefficient | 0.66 | .. | .. |
|  | Conditional R<sup>2</sup> | 0.86 | .. | .. |
| 25–34 | Intercept | -4.39 | (-8.82–0.04) | 0.085 |
|  | Time-lag (years)* | -0.11 | (-0.22– -0.01) | 0.042 |
|  | Age at sexual debut (years) | -0.13 | (-0.35–0.09) | 0.288 |
|  | Screening<sup>†</sup> | -0.7 | (-1.96–0.57) | 0.321 |
|  | Effect modification of age at sexual debut on time-lag | 0.0097 | (0.004–0.015) | 0.001 |
|  | Within-location variance | 0.07 | .. | .. |
|  | Between-location variance | 0.68 | .. | .. |
|  | Intraclass correlation coefficient | 0.91 | .. | .. |
|  | Conditional R<sup>2</sup> | 0.92 | .. | .. |
| 35–44 | Intercept | -4.32 | (-8.96–0.31) | 0.103 |
|  | Time-lag (years)* | 0 | (-0.09–0.09) | 0.985 |
|  | Age at sexual debut (years) | -0.06 | (-0.29–0.17) | 0.626 |
|  | Screening<sup>†</sup> | -0.83 | (-2.15–0.49) | 0.257 |
|  | Effect modification of age at sexual debut on time-lag | 0.0004 | (-0.004–0.005) | 0.872 |
|  | Within-location variance | 0.05 | .. | .. |
|  | Between-location variance | 0.68 | .. | .. |
|  | Intraclass correlation coefficient | 0.94 | .. | .. |
|  | Conditional R<sup>2</sup> | 0.94 | .. | .. |
| 45–54 | Intercept | -3.3 | (-8.07–1.46) | 0.215 |
|  | Time-lag (years)* | -0.06 | (-0.15–0.03) | 0.179 |
|  | Age at sexual debut (years) | -0.08 | (-0.31–0.15) | 0.508 |
|  | Screening<sup>†</sup> | -0.76 | (-2.07–0.54) | 0.294 |

|  |  |  |  |
| --- | --- | --- | --- |
| Effect modification of age at sexual debut on time-lag | 0·0025 | (-0·002–0·007) | 0·282 |
| Within-location variance | 0·04 | .. | .. |
| Between-location variance | 0·65 | .. | .. |
| Intraclass correlation coefficient | 0·94 | .. | .. |
| Conditional R<sup>2</sup> | 0·95 | .. | .. |

CI=confidence interval. HPV=human papillomavirus. \*Years elapsed between HR HPV detection and cervical cancer
incidence assessment. †Effect of organized screening with coverage >50%.

Fig 2 shows the annual incidence of cervical cancer (per 1000 HPV positive women) over a period of 14 years following HPV detection, by age group at HPV detection. Cervical cancer incidence is shown for each of the 16 locations (thin curves), as well as the model-based predictions for locations with (thick solid) and without (thick dotted) screening. For 20 to 24 year-olds, cervical cancer incidence increases by approximately one order of magnitude over the 14-year period following HR HPV detection, e.g. 0.13 (0.09-to-0.2) to 2.3 (1.6-to-3.5) per 1000 HR HPV infected women in locations without screening (see appendix p 12, Table S5 for more details). With increasing age, increases in cervical cancer risk with time since HR HPV detection are less strong, so that among 45 to 54 year-olds, cervical cancer risk plateaus between 6.6 (4.3-to-10.4) and between 5.7 (3.7-to-8.8) cancers per 1000 HR HPV infected women over the 14 years. Cervical cancer risk was higher in locations with than without screening, with this risk difference increasing with age at HPV detection (Fig 2).

**Fig 2.**
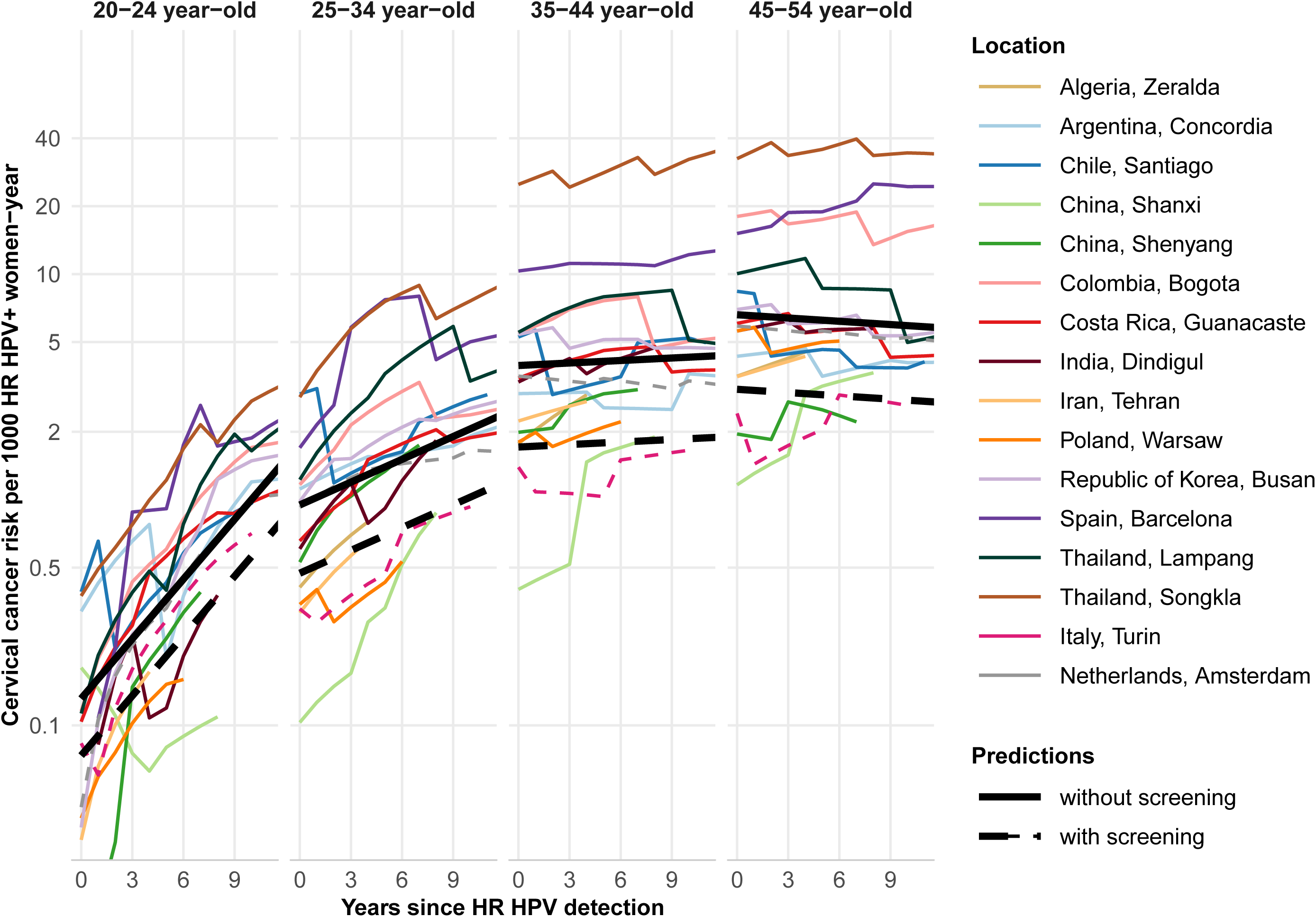
Cervical cancer incidence rates in HR HPV positive women, by years since HR HPV detection, age-group at HPV detection, screening implementation status and location. Model-based projections were drawn assuming the following average age at sexual debut in locations without and with screening, respectively, 18·3 and 16·9 years (age group 20 to 24); 19·7 and 17·6 years (age group 25–34); 20 and 17·5 (age group 35–44); and 20·3 and 17·7 (age group 45– 54). HR HPV=high-risk human papillomavirus.

Fig 3 shows estimated cervical cancer risk in the 14-year period following HR HPV detection, restricted to locations without screening, according to average sexual debut. Below 35 years-old, average age at sexual debut is an important modifier of cervical cancer risk in HPV-positive women, with earlier sexual debut being associated with a higher initial risk. For example, at age 20 to 24, the initial risk of cancer was 0.2 (0.1-to-0.3), 0.06 (0.03-to-0.1), and 0.02 (0.003-to-0.06) for average sexual debut at 17, 20, and 23 years, respectively. However, these risks converge 14 years later: 2.3 (1.4-to-3.8), 2.2 (1.1-to-4.6), and 2.2 (0.5-to-10.7), respectively. In contrast, in women with HR HPV detection at older ages, age at sexual debut was not a strong modifier of cervical cancer risk. More details about the estimated risk of cervical cancer in HPV positive women for locations with and without screening are provided in appendix pp 13-14, Table S6. Also, panels in appendix p3, Fig S2, display the match between predictions drawn using the fixed effect component of the regression models and cervical cancer incidence among HPV positive women in each location.

**Fig 3.**
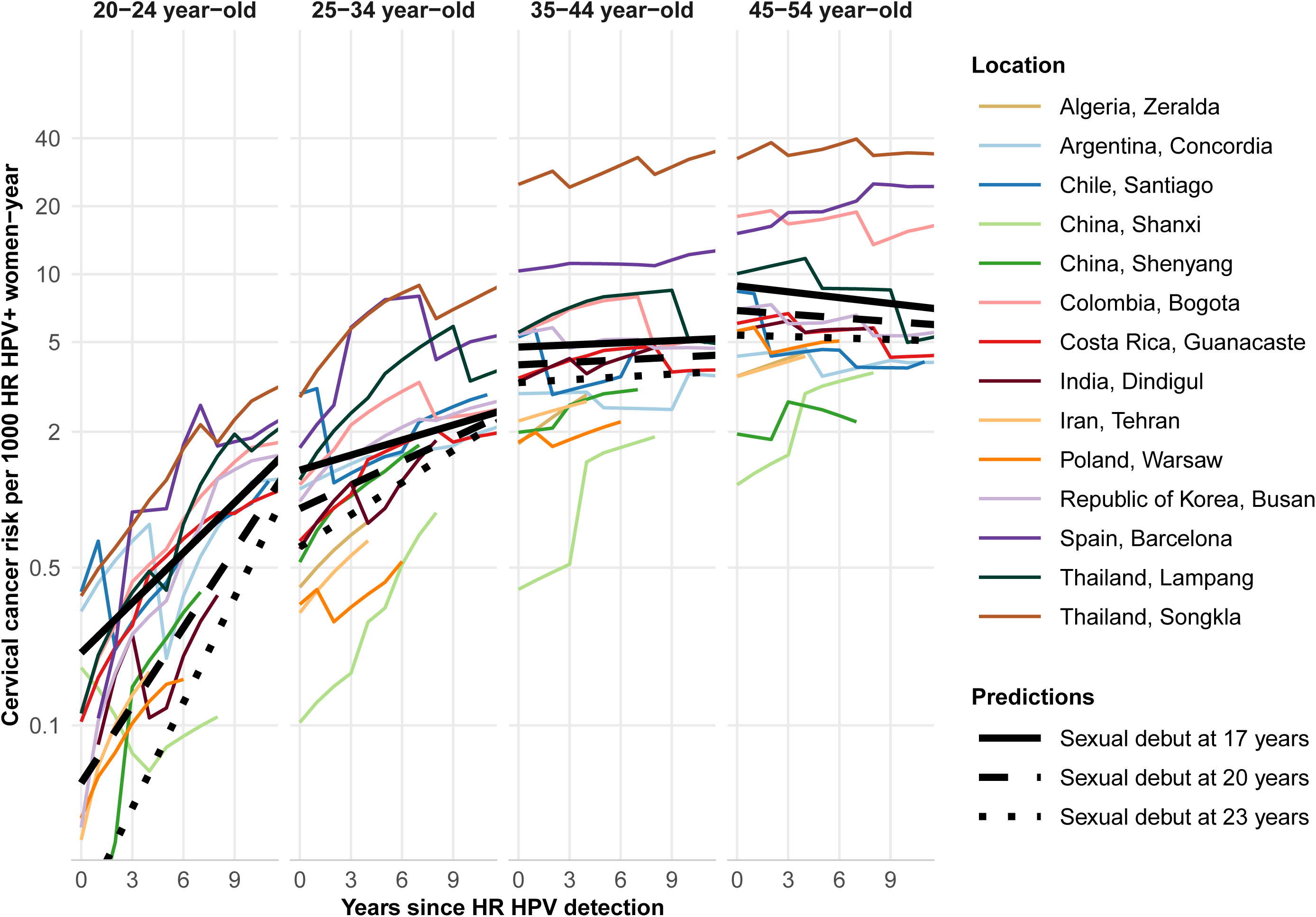
Cervical cancer incidence rates in HR HPV positive women in locations without screening, by years since HR HPV detection, age-group at HR HPV detection, average age at sexual debut, and location. HR HPV=high-risk human papillomavirus.

Table 3 shows, for IARC population-based surveys without corresponding cancer registries, and hence excluded from previous analyses, the predicted annual number and incidence of cervical cancer (with 95% PIs) in the population of the corresponding country ten years later. In all countries, the predicted annual incidence increased with age, with the lowest predicted incidence in Pakistan, ranging from 1.2 (0.5-to-2.9) to 12.6 (5.0-to-30.8) per 100 000 women, among 30-34 and 55-64 year-old women, respectively, and the highest predicted incidence in Guinea, ranging from 34 (13.5-to-87.9) to 262 (91.5-to-715.3) per 100 000 women.

**Table 3.**
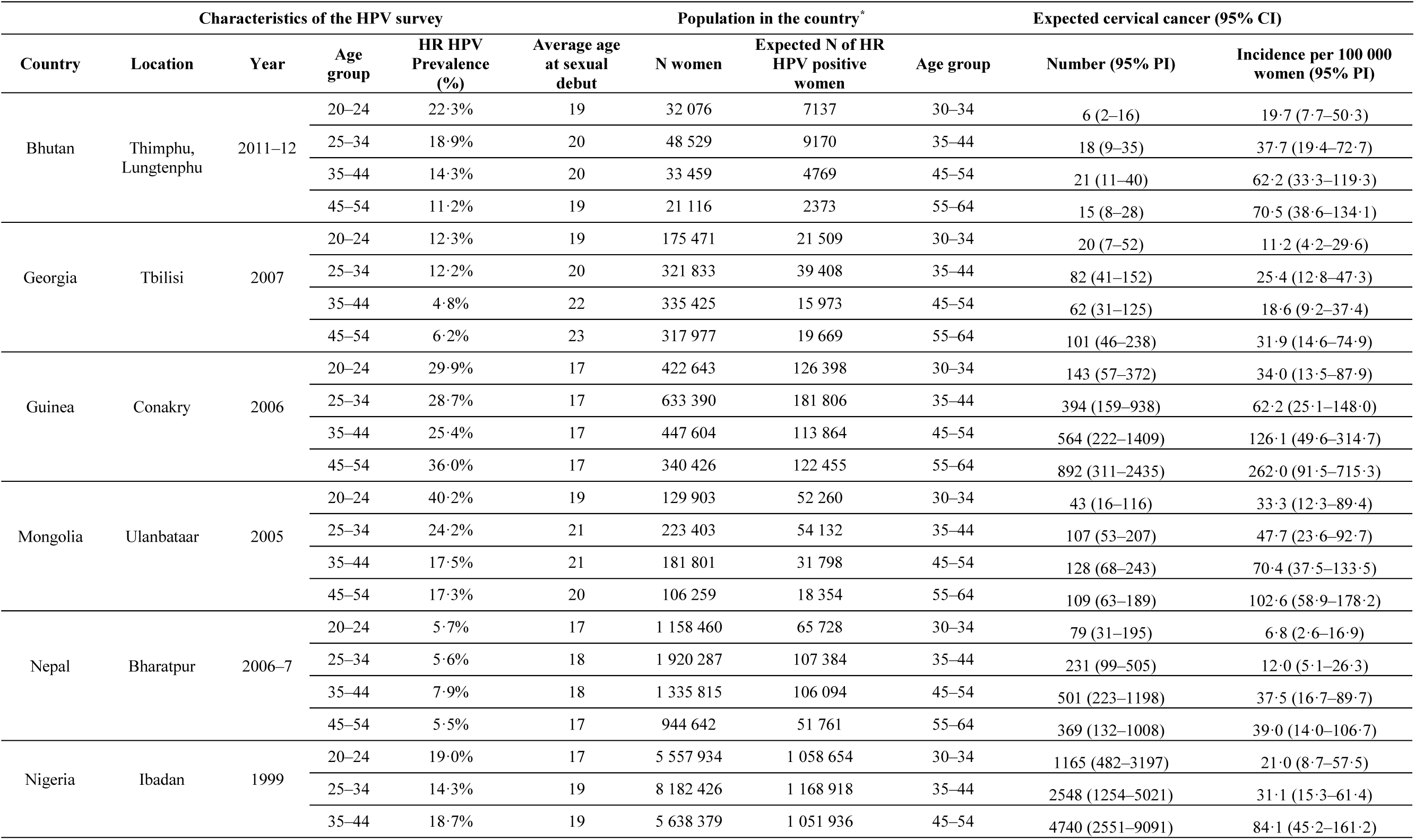
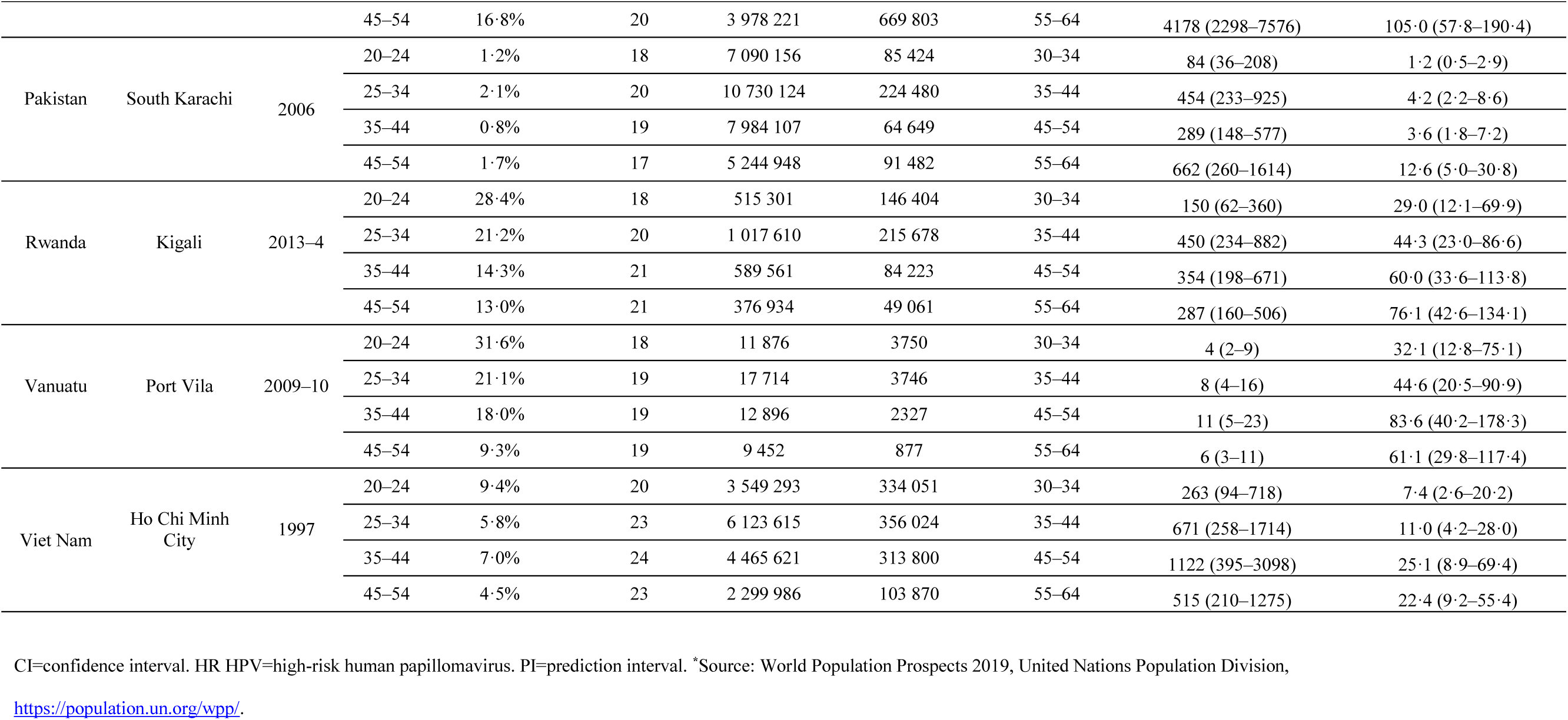
Expected annual number and incidence (per 100 000 women) of cervical cancer, ten years after the implementation of the surveys.

## Discussion

We have characterized and quantified the relationship between age-specific HPV prevalence and cervical cancer incidence in the same female birth cohorts. We found a strong and positive effect of HR HPV prevalence on cervical cancer incidence, which increased with age at HPV detection and with time-lag between HR HPV prevalence and cancer incidence assessment. In a mixed model approach, we went on to show that annual cervical cancer incidence among women with prevalent HPV infection increases with age at HPV detection and, that in women below age 35, cervical cancer risk changes by average age at first sex and increases in the 14 years following HR HPV detection. In the absence of screening, depending upon the combination of age at prevalent HR HPV detection and age at sexual debut, annual cervical cancer incidence ranged from 0.02 (0.003-to-0.06) to 8.4 (3.3-to-22.0) per 1000 HR HPV positive women. The model showed that annual cervical cancer incidence among HPV positive women was lower in screened than unscreened populations. Thus, this model allows predictions of annual cancer incidence from HPV prevalence data, in countries without cancer registration, an exercise that we performed on a set of other IARC population-based HPV surveys.

Ecologic inference has been previously used by Maucort-Boulch *et al*.^25^ and Sharma *et al*.^26^ to assess international correlation between HPV prevalence and cervical cancer incidence measured in the same time period and same age groups in 13 and 40 different locations, respectively (i.e. without accounting for the time-lag between HR HPV prevalence and cancer incidence assessment to assess the birth-cohort-specific correlation). In both analyses, HPV prevalence was a better predictor of cervical cancer at older ages, probably because prevalent HPV infections in older women are more likely to be persistent and at a higher risk of causing a cervical cancer.^27^ This hypothesis is reinforced by our finding that the association between HPV prevalence and cervical cancer incidence increased not only with age at HPV detection but also with the time-lag between HR HPV prevalence and cancer incidence measurement.

Our observations a) that cervical cancer incidence among women below 35 years with prevalent HR HPV infection is modulated by age at first sex, as a proxy of early exposure to HPV infection, and b) that cervical cancer incidence increases until approximately 45 years of age and subsequently remains constant, are consistent with data from unscreened populations^28^ and with the hypothesis that risk inflexion in middle age corresponds to a drop in circulating sex hormone levels during the perimenopausal period.^29^ Plummer et al. also observed an increase of cervical cancer risk associated with time since first sexual intercourse, while analysing a very large epidemiological data set on cervical cancer.^30^ Similarly, their risk estimates flattened approximately at age 45 years and remained constant at older ages.

We have adapted our study design to account for some typical limitations of ecological studies. First, to account for the latency between exposure and outcome (which is missed by concurrent measurement of HR HPV prevalence and cervical cancer incidence), we have assessed the two measurements in the same birth cohorts and have incorporated the time-lag between the two measurements. Second, we matched HPV and cervical cancer data from the same (or similar) location, to geographically match, as far as possible, exposure and outcome measurements. Third, we accounted for age at first sex and cervical cancer screening as a possible confounder of the ecologic relationship in the same locations between HPV prevalence and cervical cancer incidence. The effect of screening in Amsterdam and Turin on the risk of cervical cancer among HPV positive women (that was detectable with a fixed effect model - data not shown), was accounted for in the mixed effect model by the between-location variance and intraclass correlation coefficient. Analogously, the effect of other location-related determinants of cervical cancer risk given prevalent HR HPV detection, such as HIV status, reproductive behaviour factors, hormonal contraceptive use, and smoking, for which we were unable to explicitly account for, may also have been captured by the between-location variance. Consequently, our predictions of the annual incidence of cervical cancer in countries not represented in CI5 are based on average age at sexual debut, time elapsed since HPV assessment, as well as on the age-specific HR HPV prevalence assessed in each survey. The accuracy of the reported predictions relies on the assumption that the age-specific HR HPV prevalence assessed in the survey is representative of the prevalence in the whole country.

Our estimates of the risk of cervical cancer among HR HPV positive women are based upon the systematic use of the GP5+/6+ test, which is validated for clinical purposes. Hence our findings should be applicable to other population-based datasets obtained using HPV tests with a similar sensitivity, but may be less applicable to prevalence data generated with more sensitive methods.

In conclusion, using worldwide, high quality, standardized data on age-specific HPV prevalence and cervical cancer incidence, we have estimated age-specific cervical cancer incidence over time elapsed since prevalent HPV detection as a function of average age at first sex in the female population. This finding can be particularly useful to design and plan cancer control programmes in settings without cancer registries, as it allows, as exemplified above, a predictive assessment of the expected burden of cervical cancer from data collected through population-based cross-sectional HPV prevalence surveys, which are relatively easy to implement.

## Data Availability

Incidence: C15 Vol. VIII-XI (1993-2012)
Prevalence: IARC Prevalence Surveys (1993-2008)
Population: UN Population Prospects (1993-2012)
Mortality: UN Population Prospects (1993-2012)

## Contributors

RSF and IB conceived and designed the study. RSF, DG, and IB collected and analysed the data. RSF, GC, and IB drafted the manuscript. All authors contributed to the interpretation of data and approved the final manuscript.

## Funding

This work was supported by the Bill & Melinda Gates Foundation [OPP 1188709]; and the Canadian Institutes of Health Research [334069].

## Declaration of interests

We declare no competing interests.

## Acknowledgments

The authors would like to thank Professor Delphine Maucort-Boulch for her technical advice, and Doctor Silvia Franceschi, Professor Julian Peto and Doctor Guglielmo Ronco for their helpful comments. Where authors are identified as personnel of the International Agency for Research on Cancer and WHO, the authors alone are responsible for the views expressed in this article and they do not necessarily represent the decisions, policy or views of the International Agency for Research on Cancer and WHO.

## Notes

### Competing Interest Statement

The authors have declared no competing interest.

### Clinical Trial

N/A

